# A Framework to Quantify Disparities in Pharmacogenomic Treatment Concordance and Drug Response Outcomes

**DOI:** 10.1101/2025.09.29.25335264

**Authors:** Ilia Rattsev, James M. Stevenson, Casey Overby Taylor

## Abstract

Clinical PGx practice guidelines (PGx guidelines) may have limited generalizability for “marginalized” groups. We proposed the five-step Real-World Data for Genome-Guided Prescribing (ReGGRx) framework and, using All of Us research program (AoU) data, examined its ability to estimate disparities in concordance with and benefit from PGx guidelines for *CYP2C19* testing when choosing antiplatelet and antidepressant drugs. The selected measures were intended to identify disparities in avoiding drug failure independent of following PGx guidelines, the odds of avoiding drug failure with PGx concordant treatment, and the degree to which “marginalized” groups (i.e., groups underrepresented in biomedical research [UBR] and with indeterminate CYP2C19 phenotypes) benefit from PGx concordant treatment, when compared with “non-marginalized” groups (i.e., non-UBR and known CYP2C19 phenotypes). Our findings identified disparities in the antidepressant cohort with UBRs (32% of cohort) having a lower odds of avoiding drug failure. For both cohorts, a lower probability of avoiding drug failure was observed in the indeterminate phenotype group (1% of cohorts) than in the known phenotype group, indicating a need to better characterize rare or ancestry-specific risk alleles. With PGx concordant treatment, negative equal opportunity difference values suggested that the UBR group was less likely to avoid drug failure than the non-UBR group. Overall, our findings illustrate the promise of the ReGGRx framework to assess PGx guideline generalizability and produce evidence for use in drug policy decisions.

## 1. INTRODUCTION

Institutional medication use policies should consider information regarding the safety and effectiveness of drug therapy in the local patient populations they treat. This information is particularly important when considering pharmacogenomics (PGx)-guided drug policy that includes genetic variants with other clinical characteristics (age, body weight, etc.) to optimize therapeutic efficacy and minimize toxicity. However, genetic differences (e.g., risk alleles found predominantly in individuals of a certain ancestry) across ancestral groups can have clinically significant effects on drug efficacy and safety.^1^ With much of PGx research to date focusing on individuals of European ancestry,^2^ PGx evidence may have limited generalizability to non-European populations.^3^ Furthermore, the use of race as a proxy for genetic ancestry in PGx research is problematic because broad racial categories have a social, rather than biological nature and do not capture complex genetic diversity within individuals identifying with the same race. Limited understanding of PGx in non-European populations can have a detrimental effect when PGx is used clinically in diverse patient populations.^1^ When PGx tests or guidelines fail to account for genetic variants predominantly found in non-European populations, patients from other backgrounds may have their PGx results improperly interpreted or applied^4^. As a result, PGx-guided care may lack benefit or even trend toward harm for these populations^5,6^.

To generate evidence that can be used to support implementation decisions around pre-treatment PGx testing, we proposed the Real-World Data for Genome-Guided Prescribing (ReGGRx) framework and applied it to All of Us (AoU) research program data to derive measures with which to assess the potential fairness of such testing according to PGx guidelines in a real-world setting. We explored PGx guidelines regarding *CYP219* genotype assessment for antiplatelet and antidepressant therapy selection as a case study. Electronic health record (EHR) data and PGx phenotypes^7^ of AoU participants were analyzed under the assumption that they had not had genotype-guided drug therapy at the time of our study.

We considered two clinical guidelines from the Clinical Pharmacogenomics Implementation Consortium (CPIC)^8^: *CYP2C19* genotype testing for choice of antiplatelet therapy^9^ and for dosing and selection of selective serotonin reuptake inhibitor (SSRI) antidepressants.^10^ The antiplatelet clopidogrel is a prodrug that requires bioactivation into an active metabolite, which is predominantly done by CYP2C19 ^11,12^ . CPIC guidelines recommend that individuals with reduced CYP2C19 activity (CYP2C19 poor or intermediate metabolizers) avoid taking clopidogrel as an antiplatelet drug and choose an alternative P2Y12 inhibitor instead.^9,13^ The antidepressants citalopram and escitalopram are metabolized primarily by CYP2C19 into inactive metabolites^14^. CPIC guidelines recommend that CYP2C19 ultrarapid or poor metabolizers avoid taking citalopram and escitalopram for depression. Antidepressants not majorly metabolized by CYP2C19 should be considered instead, and if this is not possible, dose adjustments can be considered.^10,15^

To explore disparities in benefiting from PGx guidelines, we considered individuals from groups underrepresented in biomedical research (UBR) and with indeterminate CYP2C19 phenotypes as “marginalized” groups for comparison with “non-marginalized” groups. We specifically assessed: (1) demographic and clinical characteristics among the antiplatelet and antidepressant cohorts, (2) existing disparities in avoiding drug failure independent of whether PGx guidelines were followed, (3) the odds of avoiding drug failure when PGx guidelines were followed, and (4) the degree to which marginalized groups benefit from PGx guidelines, when compared with non-marginalized groups.

## 2. MATERIALS AND METHODS

### 2.1. Real-World Data for ReGGRx Framework

The ReGGRx framework includes five key steps applied to an AoU research program study sample to estimate disparities in concordance with and benefit from PGx guidelines. The AoU data include multiple racial subgroups^16^ and a variety of data sources,^17,18^ including EHR and whole genome sequencing (WGS) data. The selected measures are intended to identify disparities in drug failure avoidance independent of PGx concordant treatment, the odds of avoiding drug failure with PGx concordant treatment, and the degree to which “marginalized” groups (i.e., groups underrepresented in biomedical research [UBR] and with indeterminate CYP2C19 phenotypes) benefit from PGx concordant treatment, when compared with “non-marginalized” groups (i.e., non-UBR and known CYP2C19 phenotypes).

#### 2.1.1. Step 1: Cohort Selection

First, we selected AoU participants based on individual drug exposure (version 7). Of 245,388 enrolled subjects who shared their EHR and WGS data, we formed two cohorts. The first consisted of individuals with at least one record of P2Y12 inhibitor prescription *(antiplatelet cohort)*. We excluded those who had cangrelor as the only P2Y12 inhibitor on record, as it is an intravenous medication primarily used in short-term acute care scenarios. The second cohort consisted of individuals who had at least one record of an antidepressant prescription *(antidepressant cohort)*. The list of antidepressants and P2Y12 receptor antagonists was obtained by searching NIH’s RxNav data browser and was confirmed by a clinical PGx expert (J.S.) (**Supplementary Table 1**).^19^ AoU participants were excluded if they died within the study period. In addition, we excluded individuals with <30 days between their first and last medication prescriptions.

#### 2.1.2. Step 2: Determine Demographic and Clinical Characteristics

*Sociodemographic variables.* Sociodemographic variables were obtained from the *person* domain within AoU and included age, sex, gender, race, and ethnicity. Participant age at index date (date of medication initiation) was calculated from date of birth. In addition, we adapted the definition of UBR status from the AoU initiative.^20^ We excluded age from the UBR definition because older participants are well-represented in research pertaining to the medications of interest in our study. Finally, income, education, geography, disability, and healthcare access were excluded from our UBR definition because these data were unavailable.

*Comorbid conditions*. To calculate the number of comorbidities, we identified Charlson comorbidity index^21,22^ conditions recorded for study participants before the index date. We used the *comorbidiPy* library (version 0.5.0) in Python (version 3.10.12) to map ICD9 and ICD10 diagnostic codes to conditions. We then counted the Charlson comorbidities present for each participant at index date. As several conditions (acute myocardial infarction, peripheral vascular disease, and cerebrovascular disease) are parts of the outcome definition for the antiplatelet cohort, we excluded those conditions from the comorbidity count for the antiplatelet cohort.

*Pharmacogenomic phenotypes and CPIC-recommended medications.* We used the PGx phenotype calls previously annotated by Haddad at al.^23^ Out of three genes included in CPIC guidelines for SSRIs (*CYP2C19*, *CYP2B6*, and *CYP2D6*), only CYP2C19 phenotypes were available for further analysis. Based on allele functionality, CYP2C19 phenotype was captured for each participant as *ultrarapid* metabolizer, *rapid* metabolizer, *normal* metabolizer, *likely intermediate* metabolizer, *intermediate* metabolizer, *likely poor* metabolizer, or *poor* metabolizer. Participants who had a phenotype of an uncertain effect were considered CYP2C19 indeterminate.

#### 2.1.3. Step 3: Estimate Concordance with PGx Guidelines

For each AoU participant in our cohorts, we assessed the concordance of the index medication with the PGx guideline, regardless of whether one was considered when prescribing the medication. To identify drug exposures, we extracted drug name, concept id, and start date for each prescription from the AoU *drug_exposure* domain, according to our data extraction pipeline presented in **Appendix A**. After retaining only prescription records relevant to the cohort (P2Y12 inhibitors for the antiplatelet cohort and antidepressants for the antidepressant cohort), we defined the *index drug* as the first prescription record for the participant and the start date as the date recorded for the index drug.

For individuals in the antiplatelet cohort with reduced CYP2C19 function (actionable phenotypes: *likely intermediate*, *intermediate*, *likely poor*, and *poor* metabolizers), prasugrel and ticagrelor were labeled as PGx guideline concordant medications, whereas clopidogrel was labeled as a guideline discordant medication. For individuals in the antidepressant cohort with actionable phenotypes: *ultrarapid*, *likely poor*, and *poor* metabolizers, CPIC guidelines recommend considering alternatives to citalopram and escitalopram that are not primarily metabolized by CYP2C19.^10^ For this study, we considered the medication to be primarily metabolized by CYP2C19 if it had existing CPIC guidelines recommending a change in dose based on CYP2C19 phenotype. Such medications included the SSRI sertraline and tricyclic antidepressants amitriptyline, clomipramine, doxepin, imipramine, and trimipramine (see **Supplementary Table 1**).^24,25^ The algorithm to assess concordance with PGx guidelines in the antidepressant cohort is presented in **Figure 1**. We considered a medication to be PGx guideline concordant for phenotypes which CPIC does not recommend proactively changing dose or drug.

**Figure 1.**
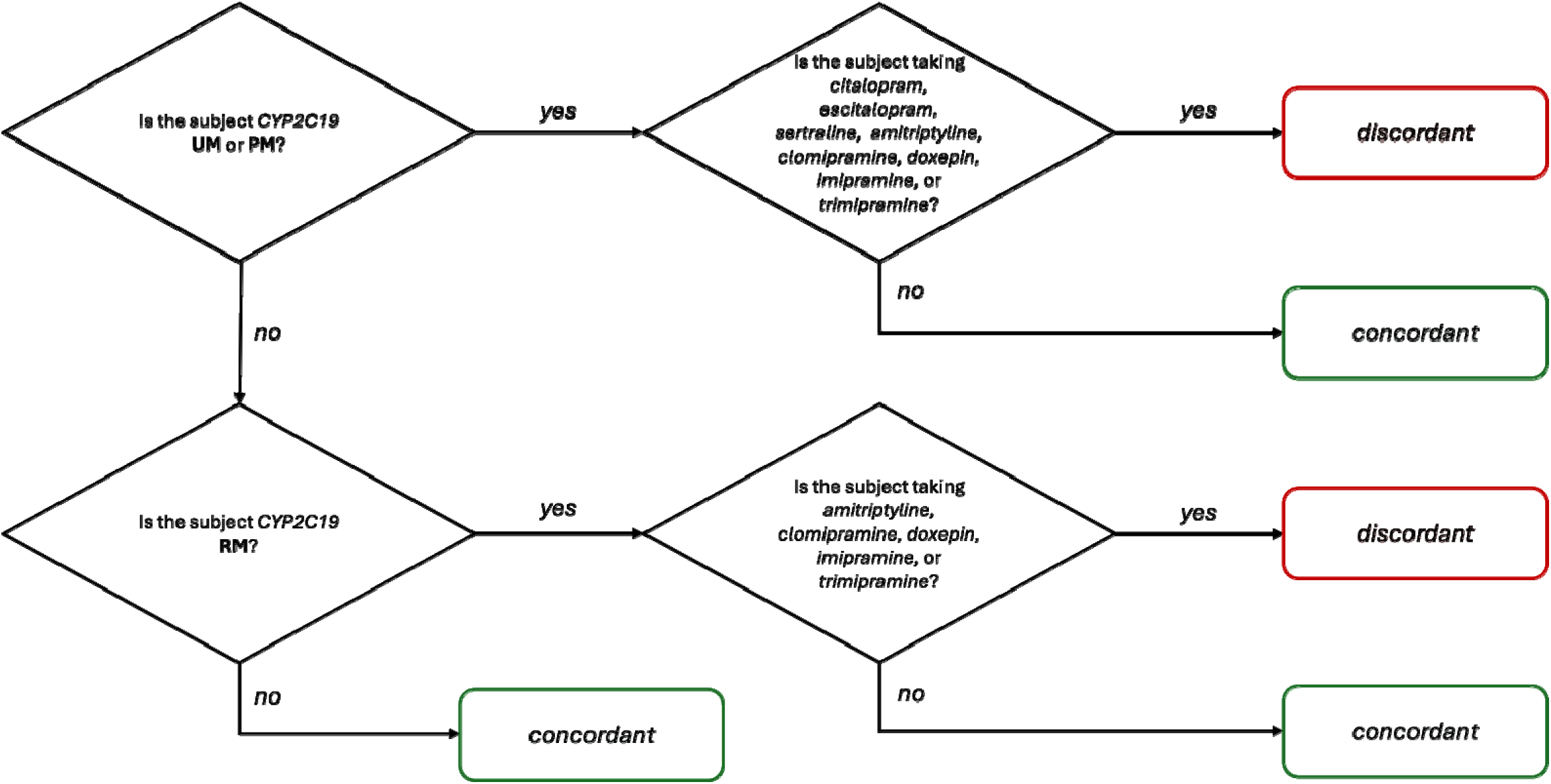
Concordance labeling algorithm for antidepressants. PM, CYP2C19 *likely poor* and *poor* metabolizer; RM, CYP2C19 *rapid* metabolizer; UM, CYP2C19 *ultrarapid* metabolizer.

#### 2.1.4. Step 4: Determine Drug Response Outcomes

The fourth step involved detecting *drug failure avoidance* as the drug response outcome of interest, given our goal to assist local pharmacy & therapeutics committees in prioritizing areas for pre-treatment PGx testing. Indicators of drug failure, summarized in **Figure 2**, were defined differently for antiplatelet and antidepressant cohorts. We limit our observation period to be one year, in alignment with the time period used by studies evaluating risk for drug failure with genotype-guided drug therapy^26,27^.

**Figure 2.**
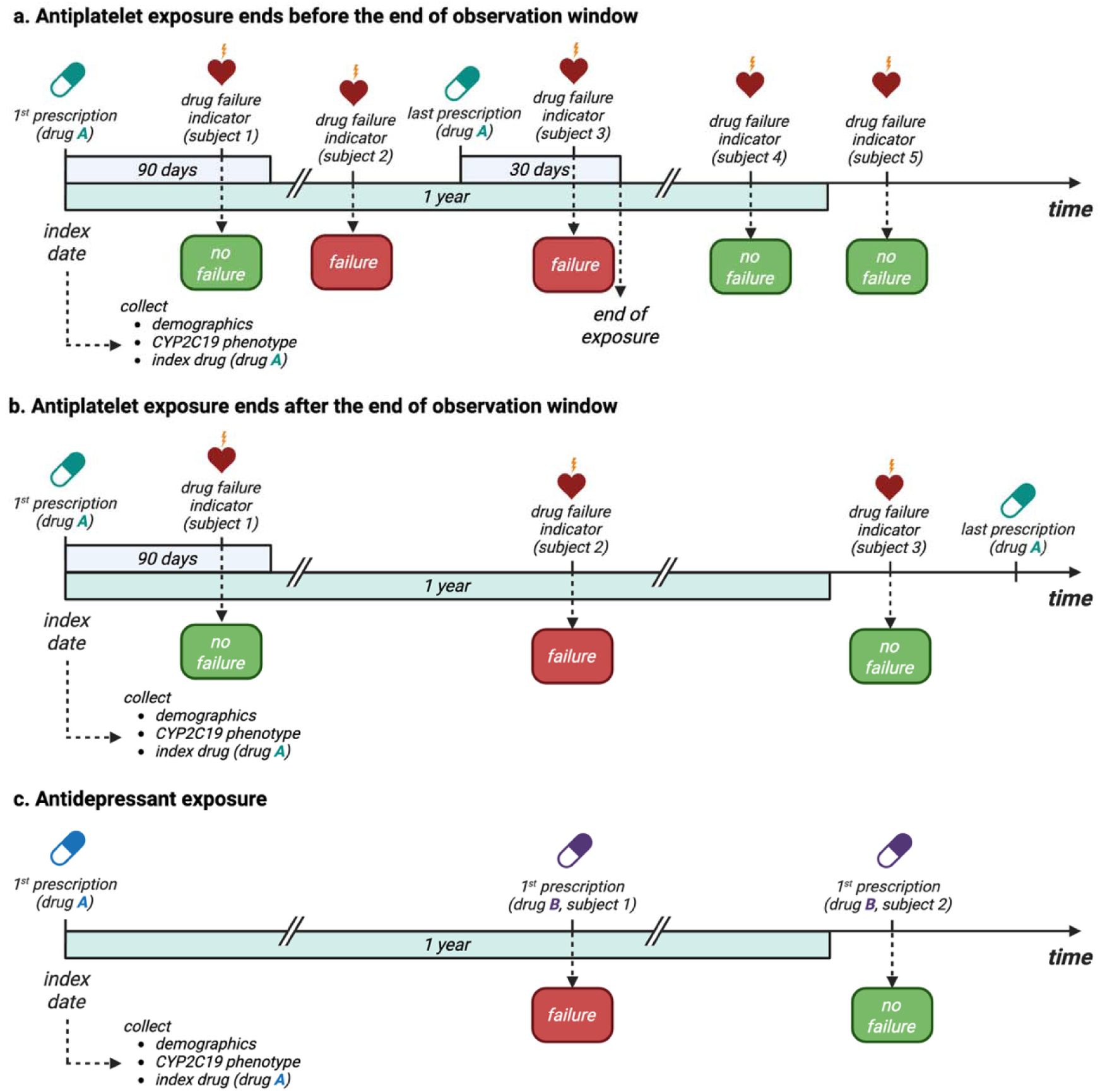
Diagram illustrating timing of measures.

For the antiplatelet cohort, a drug failure indicator was the occurrence of a condition that the medication is indicated to treat while the person was exposed to the medication. In addition, the drug failure must have occurred >= 90 days after the index date to ensure sufficient medication exposure before the drug failure event occurred. According to our definition, the medication exposure period started at the index date and ended 30 days after the start date of the last relevant prescription on record (**Figure 2a** and **Figure 2b**). The drug-relevant conditions for the antiplatelet cohort were identified based on drug labels through the FDA database.^28^ The corresponding ICD-10 codes were derived from peer-reviewed publications and are described in **Supplementary Table 2**. ICD-9 codes were converted to ICD-10 using the general equivalence mappings published by the Centers for Medicare and Medicaid Services^29^.

For the antidepressant cohort, a drug failure indicator was a medication switch (**Figure 2c**). the indications for medication prescription included depression and anxiety. We used the list of ICD-9 and ICD-10 codes provided for the phenotyping algorithms for anxiety and depression in PheKB (see **Supplementary Table 2** for the complete list of codes).^27–29^

#### 2.1.5. Step 5: Evaluate Disparities in Drug Response Outcome, Concordance with PGx Guidelines, and in Benefit from PGx Guidelines

We investigated disparities in drug failures and in concordance with PGx guidelines by calculating the odds of avoiding drug failure according to UBR status and concordance with PGx guidelines, respectively. We fit a logistic regression model to calculate odds ratios (ORs) for avoiding drug failure, adjusting for age, comorbidity count, and UBR status. These analyses were performed for individuals with any known CYP2C19 phenotype, and with a subset of the cohort that had actionable CYP2C19 phenotypes. Individuals with CYP2C19 indeterminate phenotype were excluded from this analysis due to our inability to determine the concordance status in such individuals.

We assessed benefit from PGx guidelines by comparing outcomes in UBR and indeterminate phenotype groups (“marginalized”) with those of non-UBR and non-indeterminate phenotype groups (“non-marginalized”), respectively. We also evaluated how well concordance with CPIC guidelines benefits marginalized compared with non-marginalized groups. Therefore, we calculated four fairness metrics: statistical parity difference (SPD), disparate impact ratio (DIR), average odds difference (AOD), and equal opportunity difference (EOD).^30^ We used SPD and DIR to measure how probability of the positive outcome (no drug failure) in the marginalized group compares to probability of the positive outcome in the non-marginalized group, irrespective of the PGx guideline concordance.

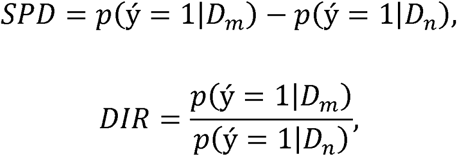

where ý=1 is the favorable outcome, D_ill_ is the marginalized group, and D_n_ is the non-marginalized group.

AOD and EOD measures were used to determine how well concordance with CPIC guidelines avoids drug failure in the marginalized and non-marginalized groups. EOD looks at the difference in true positive rate (TPR) between the groups, whereas AOD assesses the difference in terms of both TPR and false positive rate (FPR). Mathematically, these metrics are calculated as follows:

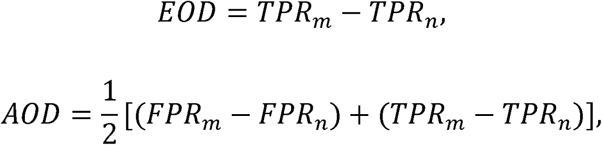

where TPR_ill_ and TPR_n_ are true positive rates for marginalized and non-marginalized groups, respectively, and FPR_ill_ and FPR_n_ are false positive rates for marginalized and non-marginalized groups, respectively (see **Appendix B**). Because CPIC guidelines cannot provide recommendations for individuals with indeterminate CYP2C19 phenotype, we were unable to calculate AOD and EOD fairness metrics for that subgroup.

Non-zero values of SPD, EOD, and AOD suggest that disparities exist in avoiding drug failure, with negative values suggesting that the marginalized group is less likely to avoid drug failure. Similarly, a DIR of 1 suggests no disparities, and values below 1 indicate a lower probability of avoiding drug failure for the marginalized group.

### 2.2. Statistical Analysis

We used a chi-squared test for independence to detect statistically significant differences between the distributions of socio-demographic characteristics in antiplatelet and antidepressant cohorts. We also performed a chi-squared test for independence to evaluate statistical significance of the differences between the distributions of phenotypes, concordance metrics, and undesirable outcomes in UBR and non-UBR groups. Statistically significant differences in distributions of continuous outcomes were analyzed by two-tailed Student’s *t*-test. We used a significance threshold of 0.05 for all tests. Bonferroni correction was used independently in the cohorts to adjust for multiple hypothesis testing. All data processing and statistical analyses were performed in AoU Researcher Workbench using Python (version 3.10.12). Statistical analysis was performed with SciPy module (version 1.11.2).^31^

## 3. RESULTS

### 3.1. Study Population

Of 70,891 AoU research program participants who satisfied our inclusion criteria, 5,622 were included in the antiplatelet cohort, and 50,291 were included in the antidepressant cohort **(Figure 3)**. The distributions of race, ethnicity, and historical representation in biomedical research were similar in the two cohorts (**Table 1**). Most participants were White (65%), not Hispanic or Latino (84%), and non-UBR (68%). Individuals self-identifying as Black or African American comprised 17% of the cohort. The antidepressant cohort was on average younger than the antiplatelet cohort (47.78 years vs 61.83 years, p<0.001), had fewer Charlson comorbidities (0.83 vs 2.17, p<0.001), and included a greater percentage of women (68% vs 37%, p<0.001).

**Figure 3.**
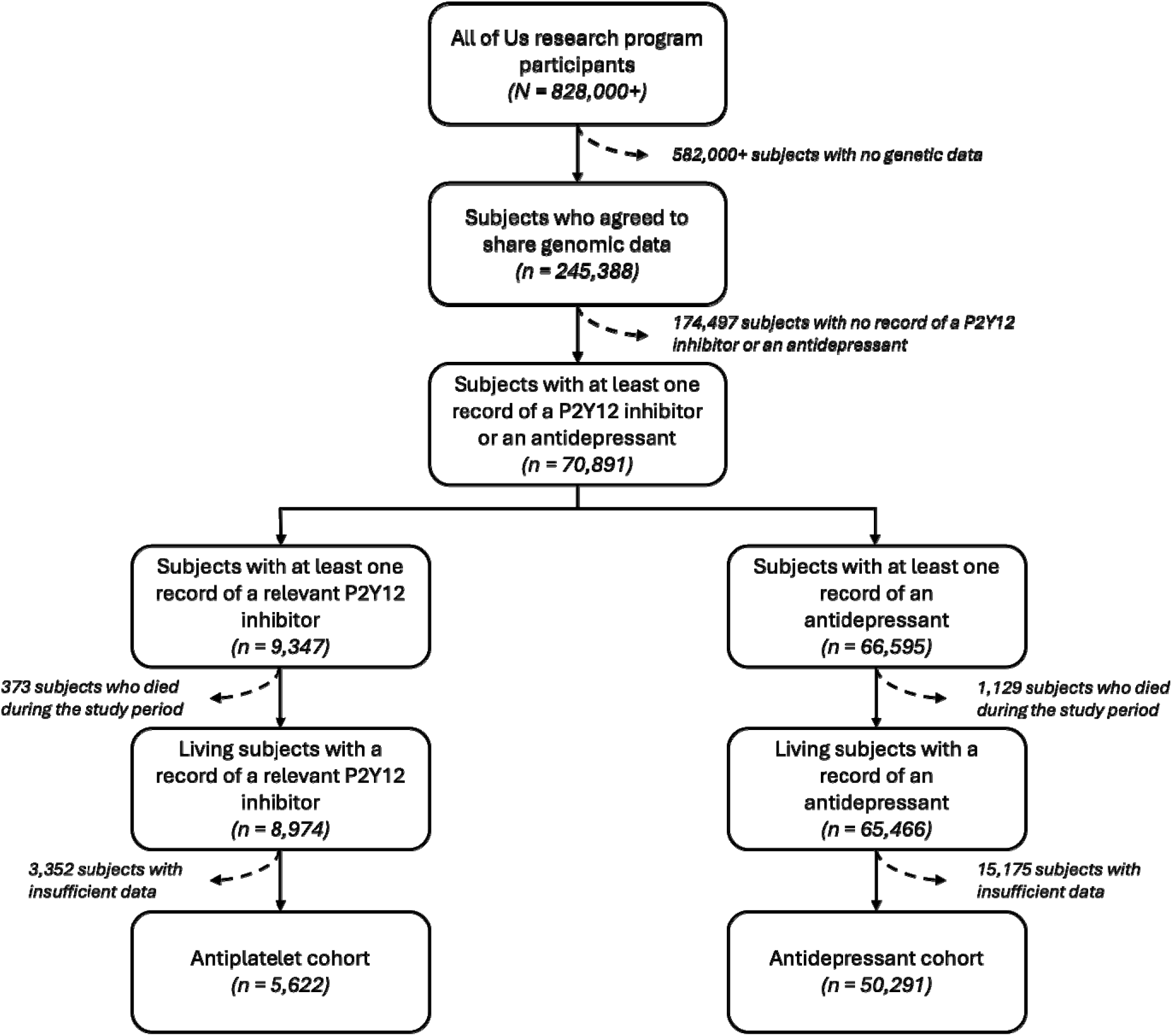
Flowchart of cohort selection.

**Table 1.**
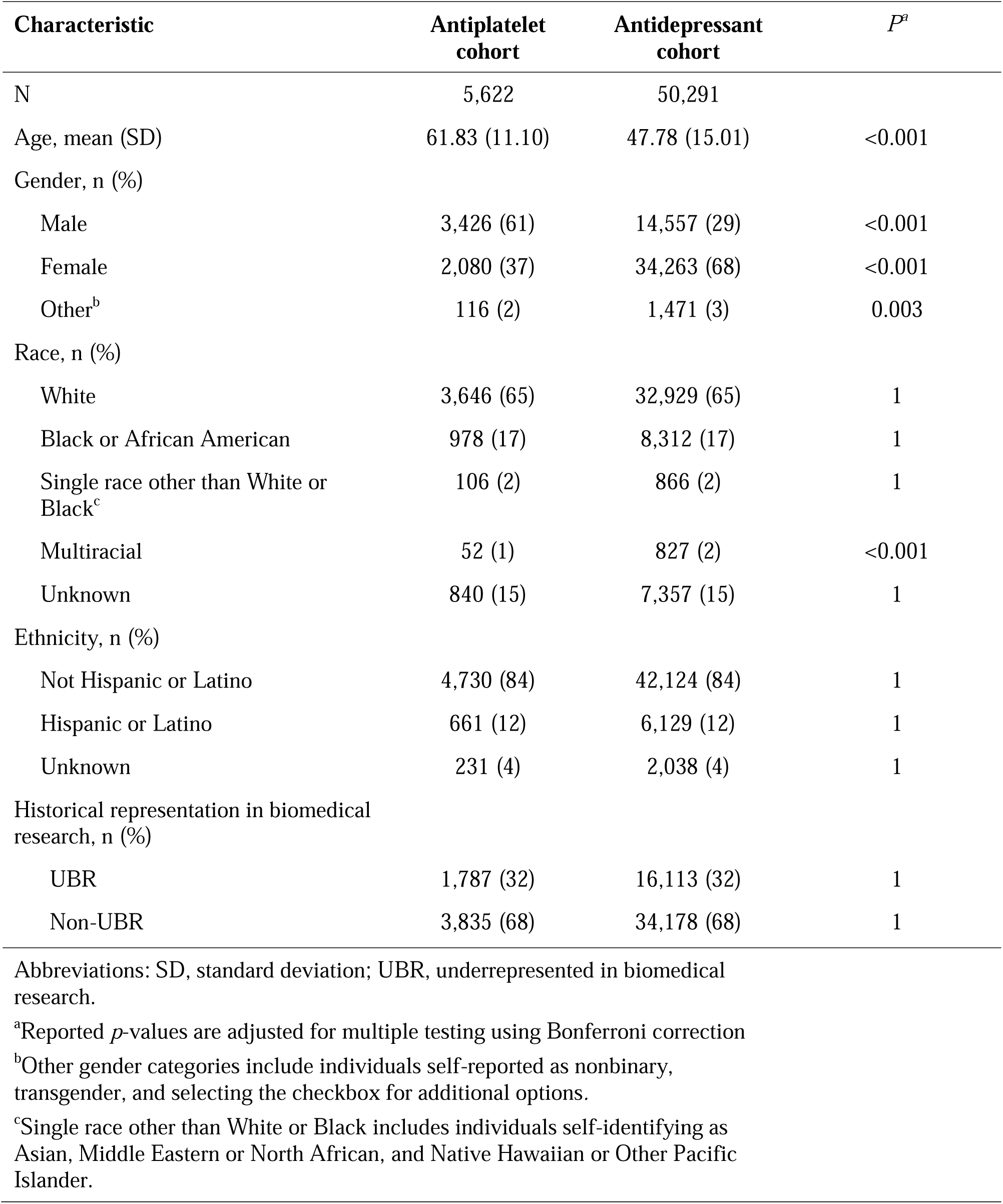
Sociodemographic and clinical characteristics of study population.

Stratification by UBR and non-UBR status revealed that UBR participants were significantly younger than non-UBR participants in both cohorts (antiplatelet cohort: 58 vs 64 years, p<0.001; antidepressant cohort: 46 vs 49 years, p<0.001; **Table 2**) and had more Charlson comorbidities (antiplatelet cohort: 2.53 vs 2.09, p<0.001; antidepressant cohort: 0.90 vs 0.80, p<0.001). CYP2C19 phenotype distributions also differed significantly. The UBR group had a higher prevalence of slower metabolizers (*poor*, *likely poor*, *intermediate*, and *likely intermediate*), whereas the non-UBR group had a larger proportion of faster metabolizers (*rapid* and *ultrarapid*). In both cohorts, the prevalence of indeterminate CYP2C19 phenotype was higher in the UBR group (2% vs <1%, p=0.002 in the antiplatelet cohort and p<0.001 in the antidepressant cohort). Significantly more non-UBR participants (75%) than UBR participants (69%) in the antiplatelet cohort had the index medication concordant with CPIC guidelines (p<0.001), but there were no significant differences in the antidepressant cohort. Finally, no significant differences in the presence of drug failure indicators were observed among UBR and non-UBR groups in either cohort.

**Table 2.** Demographic and clinical characteristics by UBR status.

| Characteristic | Antiplatelet cohort |  |  |  | Antidepressant cohort |  |  |  |
| --- | --- | --- | --- | --- | --- | --- | --- | --- |
|  | Total<br>(*N* = 5,622) | non-UBR<br>(*n* = 3,835) | UBR<br>(*n* = 1,787) | *P*<sup>*a*</sup> | Total<br>(*N* = 50,291) | non-UBR<br>(*n* = 34,178) | UBR<br>(*n* = 16,113) | *P*<sup>*a*</sup> |
| Age, y, mean (SD) | 61.83 (11.10) | 63.75 (10.67) | 57.72 (10.89) | <0.001 | 47.78 (15.01) | 48.72 (15.46) | 45.78 (13.79) | <0.001 |
| Comorbidity count, mean (SD) | 2.17 (1.76) | 2.09 (1.75) | 2.35 (1.76) | <0.001 | 0.83 (1.30) | 0.80 (1.32) | 0.90 (1.25) | <0.001 |
| CYP2C19 phenotype, n (%) |  |  |  |  |  |  |  |  |
| Normal metabolizer | 2,133 (38) | 1,457 (38) | 676 (38) | 1 | 19,737 (39) | 13,564 (40) | 6,173 (38) | 0.053 |
| Ultrarapid metabolizer | 269 (5) | 209 (5) | 60 (3) | 0.014 | 2,313 (5) | 1,716 (5) | 597 (4) | <0.001 |
| Rapid metabolizer | 1,432 (25) | 1,029 (27) | 403 (23) | 0.012 | 12,842 (26) | 9,247 (27) | 3,595 (22) | <0.001 |
| Intermediate metabolizer | 1,559 (28) | 1,017 (27) | 542 (30) | 0.059 | 13,549 (27) | 8,747 (26) | 4,802 (30) | <0.001 |
| Poor metabolizer | 165 (3) | 94 (2) | 71 (4) | 0.039 | 1,373 (3) | 704 (2) | 669 (4) | <0.001 |
| Indeterminate | 64 (1) | 29 (1) | 35 (2) | 0.002 | 477 (1) | 200 (1) | 277 (2) | <0.001 |
| Actionable<sup>b</sup> | 1,724 (31) | 1,111 (29) | 613 (34) | 0.001 | 16,528 (33) | 11,667 (34) | 4,861 (30) | <0.001 |
| Index medication, n (%) |  |  |  |  |  |  |  |  |
| Clopidogrel | 4,701 (84) | 3,183 (83) | 1,518 (85) | 1 | – | – | – | – |
| Ticagrelor | 734 (13) | 504 (13) | 230 (13) | 1 | – | – | – | – |
| Prasugrel | 187 (3) | 148 (4) | 39 (2) | 0.026 | – | – | – | – |
| Escitalopram | – | – | – | – | 4,683 (9) | 3,349 (10) | 1,334 (8) | <0.001 |
| Citalopram | – | – | – | – | 5,568 (11) | 3,954 (12) | 1,614 (10) | <0.001 |
| Other antidepressant metabolized by CYP2C19 | – | – | – | – | 12,245 (24) | 8,009 (23) | 4,236 (26) | <0.001 |
| Other antidepressant not metabolized by CYP2C19 | – | – | – | – | 27,795 (55) | 18,866 (55) | 8,929 (55) | 1 |
| Concordance, n (%)<sup>c</sup> |  |  |  |  |  |  |  |  |
| Concordant | 4,108 (73) | 2,882 (75) | 1,226 (69) | <0.001 | 46,940 (93) | 32,076 (94) | 14,864 (92) | 0.27 |
| Discordant | 1,450 (26) | 924 (24) | 526 (29) | <0.001 | 2,874 (6) | 1,902 (6) | 972 (6) | 0.27 |
| Neither | 64 (1) | 29 (1) | 35 (2) | NA | 477 (1) | 200 (1) | 277 (2) | NA |
| Drug failure indicator, n (%)<sup>d</sup> |  |  |  |  |  |  |  |  |
| Medication switch | – | – | – | – | 14,331 (28) | 9,406 (28) | 4,925 (31) | <0.001 |
| Any condition | 1,078 (19) | 724 (19) | 354 (20) | 1 | – | – | – | – |
| Angina | 160 (3) | 113 (3) | 47 (3) | 1 | – | – | – | – |
| Myocardial infarction | 384 (7) | 260 (7) | 124 (7) | 1 | – | – | – | – |
| Stroke | 534 (9) | 351 (9) | 183 (10) | 1 | – | – | – | – |

Abbreviations: SD, standard deviation; UBR, underrepresented in biomedical research.
<sup>a</sup>Reported *p*-values are adjusted for multiple testing using Bonferroni correction.
<sup>b</sup>Actionable phenotypes for antiplatelets include *intermediate* metabolizer and *poor* metabolizer. Actionable phenotypes for antidepressants include *ultrarapid* metabolizer, *rapid* metabolizer, and *poor* metabolizer. *Likely intermediate* metabolizers and *likely poor* metabolizers were combined with *intermediate* and *poor* metabolizers, respectively.
<sup>c</sup>Only individuals with a defined CYP2C19 phenotype can be labeled as concordant or discordant. Participants with CYP2C19 indeterminate phenotype were labeled as “neither.”
<sup>d</sup>Drug failure indicators for antiplatelets are clinical diagnoses of acute myocardial infarction, unstable angina, acute ischemic stroke and/or transient ischemic attack not present at the baseline..

When participants were stratified by PGx guideline concordance, we observed no differences in drug failure prevalence in the antiplatelet cohort (**Supplementary Table 3**). When stratified by condition, more concordant individuals had a myocardial infarction (8% in concordant vs 4% in discordant, p<0.001), whereas more discordant participants had a stroke (9% in discordant vs 12% in concordant, p=0.018). In the antidepressant cohort, concordant individuals were on average older than the discordant individuals (48 vs 47 years, p=0.002). No other differences were observed among concordant and discordant individuals in either cohort (**Supplementary Table 3**).

### 3.2. Existing Disparities and Avoiding Drug Failure with PGx Guideline Concordance

After participants with an indeterminate CYP2C19 phenotype were excluded, the subset of participants with an actionable CYP2C19 phenotype consisted of 1,724 individuals in the antiplatelet cohort and 16,528 individuals in the antidepressant cohort. We calculated adjusted ORs for avoiding medication failure based on UBR status and PGx guideline concordance, while adjusting for age and number of Charlson comorbidities (**Figure 4**). When we considered both actionable and non-actionable CYP2C19 phenotypes, UBR participants taking antidepressants had a lower odds of avoiding drug failure (adjusted OR: 0.90, CI: 0.86–0.94) and we saw no significant odds disparity in the antiplatelet cohort (adjusted OR: 0.94, CI: 0.81–1.10). We saw no significant effect of PGx guideline concordance on drug failure avoidance in either cohort.

**Figure 4.**
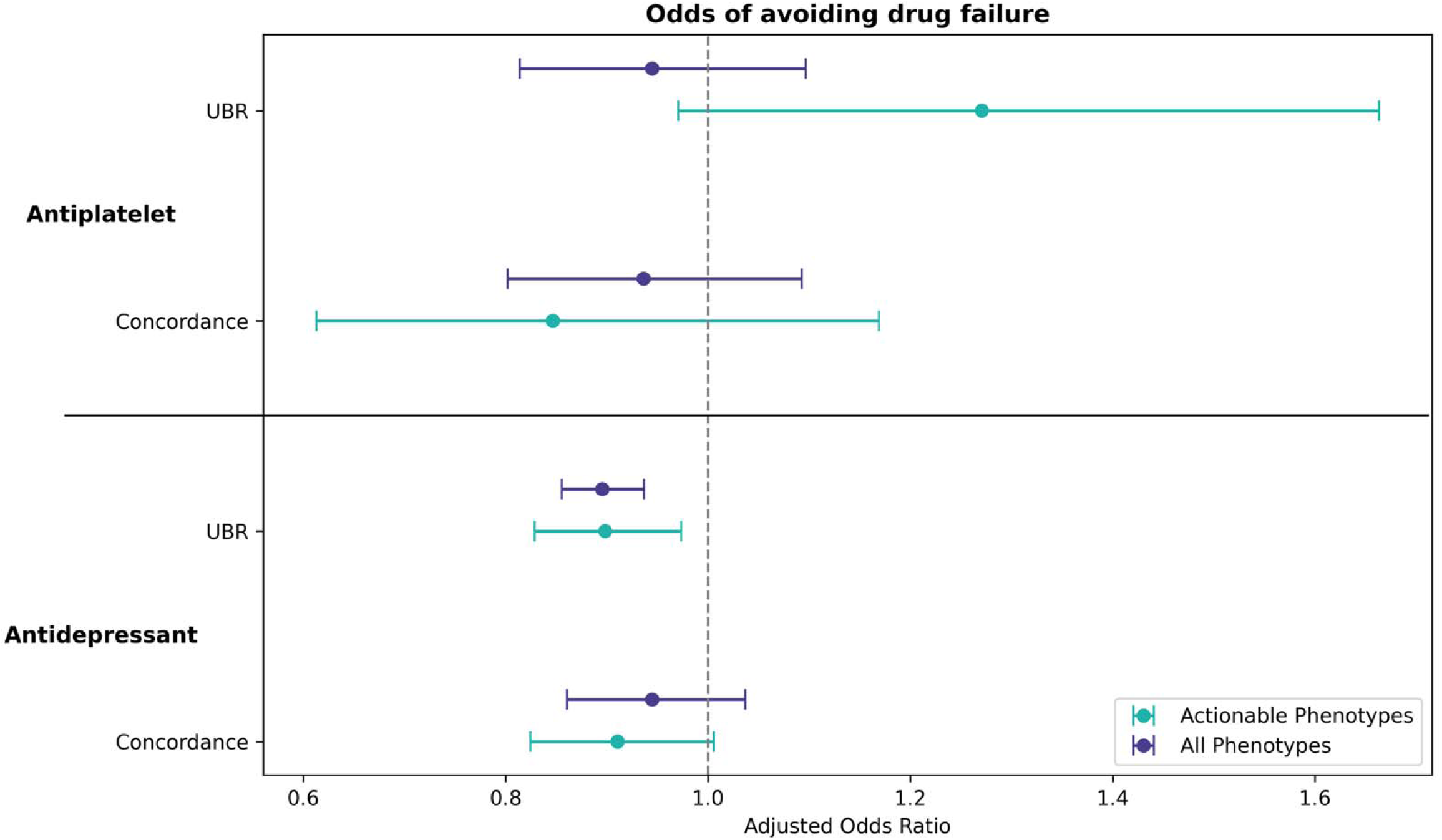
Dot plot of adjusted odds ratios (ORs) for avoiding medication failure. The plot displays ORs for avoiding medication failure adjusted for age, comorbidity count, underrepresented in biomedical research (UBR) status, and concordance of the index medication with CPIC recommendations for all individuals and those with actionable phenotypes. Error bars represent 95% confidence interval. The dashed line at OR = 1 indicates no effect. CPIC, Clinical Pharmacogenetics Implementation Consortium.

### 3.3. UBR and Indeterminate Phenotype Fairness

#### 3.3.1. Antiplatelet cohort

Among participants in the antiplatelet cohort with known CYP2C19 phenotypes, SPD and DIR metrics indicated that the probability of avoiding drug failure was slightly higher in the non-UBR group than in the UBR group (SPD: −0.01; DIR: 0.99). However, in the subgroup of individuals with actionable phenotypes both SPD and DIR suggested a lower probability of avoiding drug failure in non-UBR group (SPD: 0.04; DIR: 1.05). Among all participants, we found that the known CYP2C19 phenotype group had a higher probability of avoiding failure than the indeterminate phenotype group (SPD: −0.04; DIR: 0.95, **Figure 5**).

**Figure 5.**
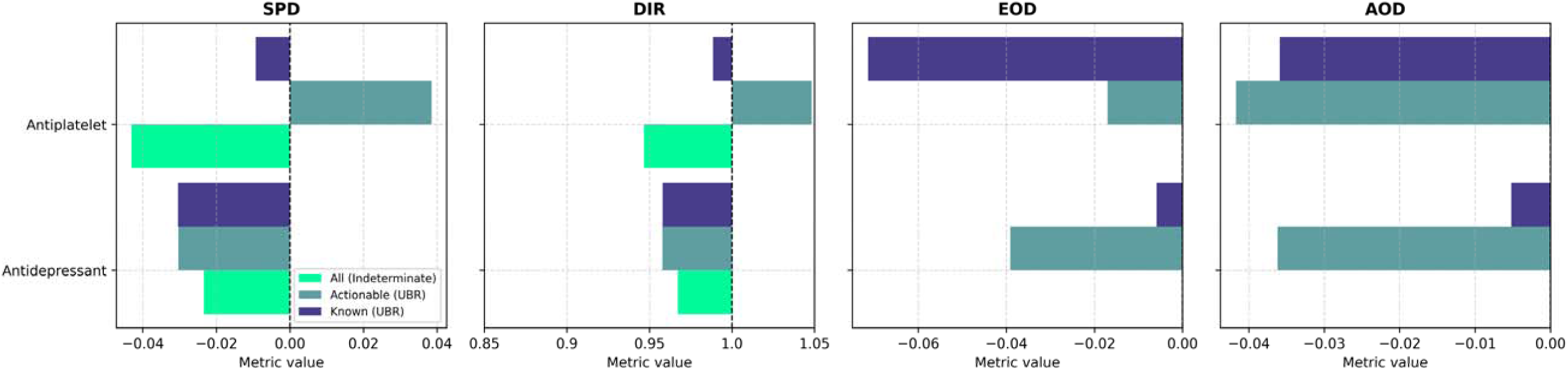
UBR and Known Phenotype Fairness metrics of CPIC guidelines. The bar plots display statistical parity difference (SPD), disparate impact ratio (DIR), equal opportunity difference (EOD), and average odds difference (AOD) of CYP2C19-related CPIC guidelines in their ability to prevent medication failure in individuals stratified by their underrepresented in biomedical research (UBR) status and CYP2C19 phenotype. In UBR fairness assessment, non-UBR was considered a non-marginalized group, and UBR was considered marginalized. In CYP2C19 phenotype fairness assessment, CYP2C19 indeterminate phenotype was considered marginalized. SPD, EOD, and AOD values of zero and a DIR value of 1 indicate perfect fairness; values below perfect fairness suggest disparity against the marginalized group, whereas values above indicate disparity against the non-marginalized group. “Known (UBR)” indicates a comparison of UBR vs non-UBR groups for those with a known CYP2C19 phenotype (N=5,558 and 49,814 in the antiplatelet and antidepressant cohorts, respectively). “Actionable (UBR)” indicates a comparison of UBR vs. non-UBR groups for those with an actionable CYP2C19 phenotype (N=1,724 and 16,528 in the antiplatelet and antidepressant cohorts, respectively). “All (Indeterminate)” indicates a comparison of the Indeterminate group vs. the group having a known CYP2C19 phenotype, for all individuals in the cohort (N=5,622 and 50,291 in the antiplatelet and antidepressant cohorts, respectively). CPIC=Clinical Pharmacogenetics Implementation Consortium.

Both EOD and AOD metrics were negative for participants taking antiplatelets (EOD: −0.07, AOD: −0.04), suggesting that non-UBR individuals benefited more from PGx guideline concordance than UBR individuals in avoiding drug failure. However, when analysis was limited to individuals with actionable phenotypes, we observed EOD values closer to zero (EOD: −0.02) and AOD values. This finding indicated that the proportion of individuals who both avoided drug failure and had PGx concordant treatment, were more similar among non-UBR and UBR groups. There was a negligible difference in AOD when limited to individuals with actionable phenotypes (AOD: −0.04 for both the known phenotype and actionable phenotype group).

#### 3.3.2. Antidepressant cohort

Among participants with known CYP2C19 phenotypes in the antidepressant cohort and in the sub-cohort with actionable phenotypes, the probability of avoiding drug failure was higher in the non-UBR group than in the UBR group (SPD: −0.03; DIR: 0.96 for participants with known phenotypes, and SPD: −0.03; DIR: 0.96 with actionable phenotypes). The cohort of participants with all phenotypes also had negative SPD and DIR values, indicating that the probability of avoiding drug failure was higher in the group with known CYP2C19 phenotypes than in the indeterminate phenotype group (SPD: −0.02; DIR: 0.97).

For the participants with a known CYP2C19 phenotype in the antidepressant cohort, we observed negative values close to zero for EOD (−0.01) and AOD (−0.01), suggesting a small disparity in avoiding drug failure with PGx guideline concordance between the UBR subgroups. However, the disparity became more apparent when we limited the analysis to individuals with actionable phenotypes; the non-UBR group benefited from the PGx guidelines more than the UBR group (EOD: −0.04, AOD: −0.04).

## 4. DISCUSSION

In this study, we presented the ReGGRx five-step framework to evaluate PGx-guided prescribing recommendations in a real-world setting and applied it to CPIC clinical guidelines for using *CYP2C19* testing to choose antiplatelet or antidepressant therapy. By applying the framework, we achieved our study objectives which enabled: (1) quantifying the representation of people that may benefit from pre-treatment PGx testing and *CYP2C19* genotype-guided prescribing, (2) clarifying where existing disparities in avoiding drug failures exist, (3) and quantifying differences in the probability of avoiding drug failures with PGx concordant treatment.

### 4.1. Representation of Individuals from Marginalized Groups, that are Discordant with PGx Guidelines, or Experienced Drug Failure in Antiplatelet and Antidepressant Cohorts

After selecting the antiplatelet and antidepressant cohorts in Step 1 of the ReGGRx framework, in Steps 2-4 enabled us to report demographic and clinical characteristics, concordance with PGx guidelines and the drug failure outcome among those cohorts (**Table 1** and **Table 2**). Those data enabled us to clarify representation of individuals from marginalized groups, that are discordant with PGx guidelines and/or have experienced a drug failure. Our findings showed a high representation of UBR individuals in both cohorts (**Table 1**, 32%, N=1,787 in the antiplatelet and 32%, N=16,113 in the antidepressant cohort). We also found a small proportion of individuals with indeterminate CYP2C19 phenotypes in the study cohorts (**Table 2**, 1%, N=64 in the antiplatelet and 1%, N=477 in the antidepressant cohort).

For most people with a known CYP2C19 phenotype, pre-treatment PGx testing would enable higher concordance with PGx guidelines, particularly for those taking antiplatelets. In the antiplatelet cohort, around 30% of individuals had a phenotype for which clopidogrel is recommended to be avoided. Given that 84% of individuals had clopidogrel as their first prescribed antiplatelet, it is unsurprising that a relatively large proportion of individuals (26%, N=1,450) were discordant with PGx guidelines. In the antidepressant cohort, 33% of individuals had a phenotype for which the PGx guideline recommends avoiding citalopram and escitalopram. Although 20% of participants were prescribed one of those medications first, only 6% (N=2,874) were discordant with the PGx guidelines. Our cohorts also had a high proportion of drug failures (19% in the antiplatelet cohort and 28% in the antidepressant cohort). These proportions are within ranges observed in other studies of antiplatelet^32,33^ and antidepressant^34,35^ drug response outcomes. For both cohorts, the large number of individuals from marginalized groups, discordant with PGx guidelines, or experiencing drug failures enabled us to clarify the potential opportunity for more pre-treatment PGx testing and genome-guided prescribing in the following steps of the ReGGRx framework.

### 4.2. Existing Disparities in Avoiding Drug Failure and Concordance with PGx Guidelines

In Step 5 of the ReGGRx framework, we identified instances where marginalized groups are less likely to avoid drug failure or to have PGx concordant treatment than non-marginalized groups. In the antidepressant cohort, we found that UBR participants had a lower odds of avoiding drug failure (**Figure 4**). Other studies have found that higher socioeconomic status is associated with higher antidepressant response^36–38^. One study of antidepressant switches among UK Biobank participants, however, found no differences according to self-reported ethnic background^38^. The AoU research program has a greater representation of UBR individuals when compared to UK Biobank and better reflects the diversity of the USA for our analyses.

When considering indeterminate and known CYP2C19 phenotype groups, a higher probability of avoiding drug failure was observed in the known phenotype group in both cohorts (**Figure 5**). This finding indicates a need to better characterize rare or ancestry-specific alleles found in UBR populations, reducing the number of individuals with indeterminate phenotypes that cannot receive PGx-guided care.

### 4.3. Avoiding Drug Failure with PGx Concordant Treatment

In Step 5 of the ReGGRx framework, we also investigated the avoidance of drug failures with PGx guideline concordance, and compared observations in marginalized and non-marginalized groups. In both the antiplatelet and antidepressant cohorts, PGx guideline concordance had no significant effect on avoiding drug failure (**Figure 4**). This contradicts several studies specifically examining citalopram, escitalopram, or clopidogrel in specific patient populations^39–41^. However, using AoU as a data source limits our ability to characterize important aspects of medication response, such as indication.

When considering UBR groups, in the antiplatelet cohort we found a disparity that became negligible when analyses were limited to individuals with an actionable phenotype (**Figure 5**). This finding is consistent with existing literature showing a lack of racial disparity in genotype-guided antiplatelet prescribing after pre-treatment PGx testing^42^. In the antidepressant cohort, however, a small disparity in avoiding drug failure with PGx guideline concordance became more apparent when limiting analyses to those with actionable phenotypes (**Figure 5**).

This finding highlights a limitation of our analyses, which were limited to CYP2C19 phenotypes when the PGx guideline specifies also considering CYP2D6 and CTP2B6. Many participants in our cohort with a drug failure despite PGx guideline concordance, would not be labeled concordant if CYP2D6 phenotypes were also considered. Access to phenotypes of all genes relevant to the PGx guideline beyond *CYP2C19* would address metabolic variations that can differ significantly by ancestry^43^. Thus, these findings can inform more equitable drug policy by prioritizing assessments of both CYP2D6, CYP2B6 and CYP2C19 phenotypes, rather than CYP2C19 alone when prescribing antidepressants.

## 5. Limitations and Considerations for Applying the ReGGRx Framework

It is important to acknowledge the limitations of our study. First, because we could not determine cause of death, we excluded individuals who died during the study period, which may have introduced immortal time bias. However, deceased individuals represented <1% of the original cohort and are unlikely to have significantly affected the findings. Second, we identified conditions indicative of drug failure in the antiplatelet cohort by using ICD diagnostic codes, which may not comprehensively capture the phenotypes of interest. To mitigate the biases that may have been introduced with this strategy, we used peer-reviewed publications and established phenotyping algorithms while compiling the ICD codes. In addition, our definition of drug failure may confound the relation between PGx guideline concordance and the drug failure outcome because a subset of people will be labeled as having a drug failure when in fact the ICD codes during follow-up refer to the index event. Antidepressant medications may be switched for reasons beyond lack of efficacy or side effects, for example, drug-drug interactions with a newly prescribed drug. Even so, it can be informative to evaluate differences in the presence of drug failure indicators among groups. Alternative strategies to infer drug failure can be readily used with the ReGGRx framework. Finally, our dataset did not contain drug dosing details or PGx phenotype data for CYP2D6 and CYP2B6 at the time of analysis, limiting our antidepressant case study to only CYP2C19-related guidelines to avoid a drug. Future work is needed to more comprehensively assess PGx guidelines for prescribing antidepressants when drug dosing details and CYP2D6 and CYP2B6 phenotypes are also known. Despite the known limitations, we believe that our framework can be applied to a wide range of PGx guidelines, medication classes, and outcomes of interest. In our case studies, we evaluated the ability of PGx guidelines to avoid drug failure when CYP2C19 phenotype is known. Other indicators of drug failure might also be considered, such as the presence of specific side effects, depending on the scenario. The framework can also be expanded to cover phenotypes of multiple pharmacogenes.

## 6. CONCLUSION

We developed the ReGGRx framework to generate real-world evidence of disparities in PGx guideline concordance of prescribed medications and in avoiding drug failures, and the generalizability of PGx guidelines for marginalized groups. We demonstrated its use to assess prescribing and treatment failure outcomes by CYP2C19 phenotype in AoU research program participants receiving antiplatelet and antidepressant medications. In summary we found disparities affecting marginalized groups present in both cohorts. Use of the ReGGRx framework enabled generating evidence to encourage future research to characterize PGx alleles found in UBR populations, clarified PGx guideline generalizability for the UBR group, and highlighted opportunities for genotype-guided prescribing to benefit more people.

## Data Availability

This study used data from the All of Us Research Program's Controlled Tier Dataset version 7, available to authorized users on the Researcher Workbench.

https://workbench.researchallofus.org/

## ACKNOWLEDGEMENTS

We thank Dr. Philip Empey (University of Pittsburgh) and *All of Us* Research Program participants for helping to make this work possible. This study was supported by NIH grant R21MD19100 awarded to CO Taylor.

## AUTHOR CONTRIBUTIONS

COT and JMS conceptualized the project. IR and COT designed the study. IR developed methods for the analysis of the data. IR prepared the tables and figures. IR, JMS, and COT wrote and reviewed the manuscript.

## COMPTETING INTERESTS

All authors declare that they have no conflicts of interest.

## DATA AVAILABILITY

This study used data from the *All of Us* Research Program’s Controlled Tier Dataset version 7, available to authorized users on the Researcher Workbench.

## REFERENCES

1. Magavern EF, Gurdasani D, Ng FL, Lee SSJ. Health equality, race and pharmacogenomics. Br J Clin Pharmacol. 2022;88(1):27–33. 10.1111/bcp.14983

2. Gurdasani D, Barroso I, Zeggini E, Sandhu MS. Genomics of disease risk in globally diverse populations. Nat Rev Genet. 2019;20(9):520–535. doi:10.1038/s41576-019-0144-0

3. Ortega VE, Meyers DA. Pharmacogenetics: Implications of race and ethnicity on&#xa0;defining genetic profiles for personalized medicine. Journal of Allergy and Clinical Immunology. 2014;133(1):16–26. doi:10.1016/j.jaci.2013.10.040

4. Duconge J, Cadilla CL, Seip RL, Ruaño G. Why Admixture Matters in Genetically-guided Therapy: Missed Targets in the COAG and EU-PACT Trials. P R Health Sci J. 2015;34(3):175. Accessed July 30, 2025. https://pmc.ncbi.nlm.nih.gov/articles/PMC4770896/

5. Kimmel SE, French B, Kasner SE, et al. A Pharmacogenetic versus a Clinical Algorithm for Warfarin Dosing. N Engl J Med. 2013;369(24):2283. doi:10.1056/NEJMOA1310669

6. Drozda K, Wong S, Patel SR, et al. Poor Warfarin Dose Prediction with Pharmacogenetic Algorithms that Exclude Genotypes Important for African Americans. Pharmacogenet Genomics. 2015;25(2):73. doi:10.1097/FPC.0000000000000108

7. Haddad A, Radhakrishnan A, McGee S, et al. Frequency of pharmacogenomic variation and medication exposures among All of Us Participants. medRxiv. Published online June 13, 2024:2024.06.12.24304664. doi:10.1101/2024.06.12.24304664

8. Relling M V, Klein TE. CPIC: Clinical Pharmacogenetics Implementation Consortium of the Pharmacogenomics Research Network. Clin Pharmacol Ther. 2011;89(3):464–467. 10.1038/clpt.2010.279

9. Lee CR, Luzum JA, Sangkuhl K, et al. Clinical Pharmacogenetics Implementation Consortium Guideline for CYP2C19 Genotype and Clopidogrel Therapy: 2022 Update. Clin Pharmacol Ther. 2022;112(5):959–967. doi:10.1002/CPT.2526

10. Bousman CA, Stevenson JM, Ramsey LB, et al. Clinical Pharmacogenetics Implementation Consortium (CPIC) Guideline for CYP2D6, CYP2C19, CYP2B6, SLC6A4, and HTR2A Genotypes and Serotonin Reuptake Inhibitor Antidepressants. Clin Pharmacol Ther. 2023;114(1):51–68. doi:10.1002/CPT.2903

11. Caplain H, Donat F, Gaud C, Necciari J. Pharmacokinetics of clopidogrel. Semin Thromb Hemost. 1999;25 Suppl 2:25–28. http://europepmc.org/abstract/MED/10440419

12. Savi J. M.; Uzabiaga M. F.; Combalbert J.; Picard C.; Maffrand J. P.; Pascal M.; Herbert J. M. P; P. Identification and Biological Activity of the Active Metabolite of Clopidogrel. Thromb Haemost. 2000;84(11):891–896. doi:10.1055/s-0037-1614133

13. Scott SA, Sangkuhl K, Gardner EE, et al. Clinical Pharmacogenetics Implementation Consortium Guidelines for Cytochrome P450–2C19 (CYP2C19) Genotype and Clopidogrel Therapy. Clin Pharmacol Ther. 2011;90(2):328–332. 10.1038/clpt.2011.132

14. Yu BN, Chen GL, He N, et al. PHARMACOKINETICS OF CITALOPRAM IN RELATION TO GENETIC POLYMORPHISM OF CYP2C19. Drug Metabolism and Disposition. 2003;31(10):1255–1259. doi:10.1124/dmd.31.10.1255

15. Hicks JK, Bishop JR, Sangkuhl K, et al. Clinical Pharmacogenetics Implementation Consortium (CPIC) Guideline for CYP2D6 and CYP2C19 Genotypes and Dosing of Selective Serotonin Reuptake Inhibitors. Clin Pharmacol Ther. 2015;98(2):127–134. 10.1002/cpt.147

16. Kathiresan N, Cho SMJ, Bhattacharya R, Truong B, Hornsby W, Natarajan P. Representation of Race and Ethnicity in the Contemporary US Health Cohort All of Us Research Program. JAMA Cardiol. 2023;8(9):859–864. doi:10.1001/jamacardio.2023.2411

17. JC D, JL R, DB G, et al. The “All of Us” Research Program. N Engl J Med. 2019;381(7):668–676. doi:10.1056/NEJMSR1809937

18. Venner E, Muzny D, Smith JD, et al. Whole-genome sequencing as an investigational device for return of hereditary disease risk and pharmacogenomic results as part of the All of Us Research Program. Genome Med. 2022;14(1):34. doi:10.1186/s13073-022-01031-z

19. Zeng K, Bodenreider O, Kilbourne J, Nelson S. RxNav: A Web Service for Standard Drug Information. AMIA Annual Symposium Proceedings. 2006;2006:1156. Accessed October 10, 2024. /pmc/articles/PMC1839335/

20. Mapes BM, Foster CS, Kusnoor S V., et al. Diversity and inclusion for the All of Us research program: A scoping review. PLoS One. 2020;15(7):e0234962. doi:10.1371/JOURNAL.PONE.0234962

21. Charlson ME, Pompei P, Ales KL, MacKenzie CR. A new method of classifying prognostic comorbidity in longitudinal studies: development and validation. J Chronic Dis. 1987;40(5):373–383. doi:10.1016/0021-9681(87)90171-8

22. Gasparini A. comorbidity: An R package for computing comorbidity scores. J Open Source Softw. 2018;3(23):648. doi:10.21105/JOSS.00648

23. Haddad A, Radhakrishnan A, McGee S, et al. Frequency of pharmacogenomic variation and medication exposures among All of Us Participants. medRxiv. Published online June 13, 2024. doi:10.1101/2024.06.12.24304664

24. Whirl-Carrillo M, McDonagh EM, Hebert JM, et al. Pharmacogenomics Knowledge for Personalized Medicine. Clin Pharmacol Ther. 2012;92(4):414–417. 10.1038/clpt.2012.96

25. Whirl-Carrillo M, Huddart R, Gong L, et al. An Evidence-Based Framework for Evaluating Pharmacogenomics Knowledge for Personalized Medicine. Clin Pharmacol Ther. 2021;110(3):563–572. 10.1002/cpt.2350

26. Pereira NL, Farkouh ME, So D, et al. Effect of Genotype-Guided Oral P2Y12 Inhibitor Selection vs Conventional Clopidogrel Therapy on Ischemic Outcomes after Percutaneous Coronary Intervention: The TAILOR-PCI Randomized Clinical Trial. JAMA - Journal of the American Medical Association. 2020;324(8):761–771. doi:10.1001/JAMA.2020.12443

27. Cavallari LH, Lee CR, Beitelshees AL, et al. Multisite Investigation of Outcomes With Implementation of CYP2C19 Genotype-Guided Antiplatelet Therapy After Percutaneous Coronary Intervention. JACC Cardiovasc Interv. 2018;11(2):181–191. doi:10.1016/J.JCIN.2017.07.022

28. FDA. FDA-Approved Drugs. Accessed March 3, 2025. https://www.fda.gov/drugsatfda

29. General Equivalence Mappings - ICD-9-CM to and from ICD-10-CM and ICD-10-PCS. https://www.cms.gov/medicare/coding/icd10/downloads/icd-10_gem_fact_sheet.pdf (Accessed July 31, 2025).

30. Bellamy RKE, Dey K, Hind M, et al. AI Fairness 360: An Extensible Toolkit for Detecting, Understanding, and Mitigating Unwanted Algorithmic Bias. Published online October 3, 2018. http://arxiv.org/abs/1810.01943

31. Virtanen P, Gommers R, Oliphant TE, et al. SciPy 1.0: fundamental algorithms for scientific computing in Python. Nature Methods 2020 17:3. 2020;17(3):261-272. doi:10.1038/s41592-019-0686-2

32. Cavallari LH, Lee CR, Beitelshees AL, et al. Multisite Investigation of Outcomes With Implementation of CYP2C19 Genotype-Guided Antiplatelet Therapy After Percutaneous Coronary Intervention. JACC Cardiovasc Interv. 2018;11(2):181–191. doi:10.1016/j.jcin.2017.07.022

33. Pereira NL, Farkouh ME, So D, et al. Effect of Genotype-Guided Oral P2Y12 Inhibitor Selection vs Conventional Clopidogrel Therapy on Ischemic Outcomes after Percutaneous Coronary Intervention: The TAILOR-PCI Randomized Clinical Trial. JAMA - Journal of the American Medical Association. 2020;324(8):761–771. doi:10.1001/JAMA.2020.12443

34. Ball S, Classi P, Dennehy EB. What happens next?: a claims database study of second-line pharmacotherapy in patients with major depressive disorder (MDD) who initiate selective serotonin reuptake inhibitor (SSRI) treatment. Ann Gen Psychiatry. 2014;13(1):8. doi:10.1186/1744-859X-13-8

35. Zhdanava M, Pilon D, Ghelerter I, et al. The Prevalence and National Burden of Treatment-Resistant Depression and Major Depressive Disorder in the United States. Journal of Clinical Psychiatry. 2021;82(2). doi:10.4088/JCP.20M13699

36. Sundell KA, Waern M, Petzold M, Gissler M. Socio-economic determinants of early discontinuation of anti-depressant treatment in young adults. Eur J Public Health. 2013;23(3):433–440. doi:10.1093/EURPUB/CKR137

37. Elwadhi D, Cohen A. Social inequalities in antidepressant treatment outcomes: a systematic review. Social Psychiatry and Psychiatric Epidemiology 2020 55:10. 2020;55(10):1241–1259. doi:10.1007/S00127-020-01918-5

38. Lo CWH, Gillett AC, Iveson MH, et al. Antidepressant Switching as a Proxy Phenotype for Drug Nonresponse: Investigating Clinical, Demographic, and Genetic Characteristics. Biological Psychiatry Global Open Science. 2025;5(4):100502. doi:10.1016/J.BPSGOS.2025.100502

39. Cavallari LH, Lee CR, Beitelshees AL, et al. Multi-site Investigation of Outcomes with Implementation of CYP2C19 Genotype-Guided Antiplatelet Therapy after Percutaneous Coronary Intervention. JACC Cardiovasc Interv. 2017;11(2):181. doi:10.1016/J.JCIN.2017.07.022

40. Aldrich SL, Poweleit EA, Prows CA, Martin LJ, Strawn JR, Ramsey LB. Influence of CYP2C19 Metabolizer Status on Escitalopram/Citalopram Tolerability and Response in Youth With Anxiety and Depressive Disorders. Front Pharmacol. 2019;10(FEB):99. doi:10.3389/FPHAR.2019.00099

41. Jukić MM, Haslemo T, Molden E, Ingelman-Sundberg M. Impact of CYP2C19 genotype on escitalopram exposure and therapeutic failure: A retrospective study based on 2,087 patients. American Journal of Psychiatry. 2018;175(5):463–470. doi:10.1176/APPI.AJP.2017.17050550/ASSET/IMAGES/LARGE/APPI.AJP.2017.17050 550F2.JPEG

42. Cavallari LH, Limdi NA, Beitelshees AL, et al. Evaluation of Potential Racial Disparities in CYP2C19-Guided P2Y12 Inhibitor Prescribing after Percutaneous Coronary Intervention. Clin Pharmacol Ther. 2022;113(3):615. doi:10.1002/CPT.2776

43. Koopmans AB, Braakman MH, Vinkers DJ, Hoek HW, van Harten PN. Meta-analysis of probability estimates of worldwide variation of CYP2D6 and CYP2C19. Transl Psychiatry. 2021;11(1):1–16. doi:10.1038/S41398-020-01129-1;SUBJMETA=2489,420,648,692,706;KWRD=CLINICAL+GENETICS,SCIENTIFIC+COMMUNITY

